# Abdominal Adiposity and Lacunar Stroke: A Mendelian Randomization Mediation Study

**DOI:** 10.64898/2026.09.02.26362110

**Authors:** Guo Mengmeng, Cong Lu, Liu Zunjing

## Abstract

**Background:** Lacunar stroke accounts for approximately 25% of ischaemic strokes and is linked to metabolic and vascular risk factors. Abdominal adiposity, measured as waist-to-hip ratio adjusted for BMI (WHRadjBMI), is a heritable predictor of cerebrovascular disease independent of overall adiposity, but its biological mediators remain unclear. We performed a two-sample Mendelian randomization (MR) mediation analysis of 16 candidate biomarkers.

**Methods:** Summary-level GWAS data were obtained from the IEU Open GWAS and EBI GWAS Catalog for WHRadjBMI (exposure), 16 mediators, and lacunar stroke (outcome). Instruments met genome-wide significance (P < 5 × 10⁻⁸), LD clumping (r² < 0.001, 10,000 kb; 1000 Genomes European panel), and F-statistic ≥ 10. Inverse variance weighted regression was the primary analysis, with MR-Egger, weighted median, and MR-PRESSO sensitivity analyses. Mediation was quantified using the product-of-coefficients method with delta-method standard errors.

**Results:** Systolic blood pressure showed the largest mediation proportion (29.6%; 95% CI, 12.7%–46.5%), followed by diastolic blood pressure (29.2%; 12.1%–46.3%) and glycated hemoglobin (23.0%; −11.2% to 57.2%). Other mediators included triglycerides (10.8%), fasting insulin (9.9%), blood glucose (7.0%), HDL cholesterol (3.2%), and LDL cholesterol (1.1%). Sensitivity analyses were generally consistent, though MR-Egger intercepts suggested directional pleiotropy for glycoprotein acetyls and CRP (Path A only). Because mediators are intercorrelated, proportions cannot be summed; multivariable MR is needed for joint effects. The HbA1c estimate was imprecise and requires cautious interpretation.

**Conclusions:** Blood pressure, and possibly glycemic control, are the leading mediators linking abdominal adiposity to lacunar stroke. These findings support prioritizing blood pressure management—and potentially glycemic control—to reduce small vessel cerebrovascular disease in individuals with elevated WHRadjBMI.

**Trial registration:** Not applicable.

Graphical Abstract

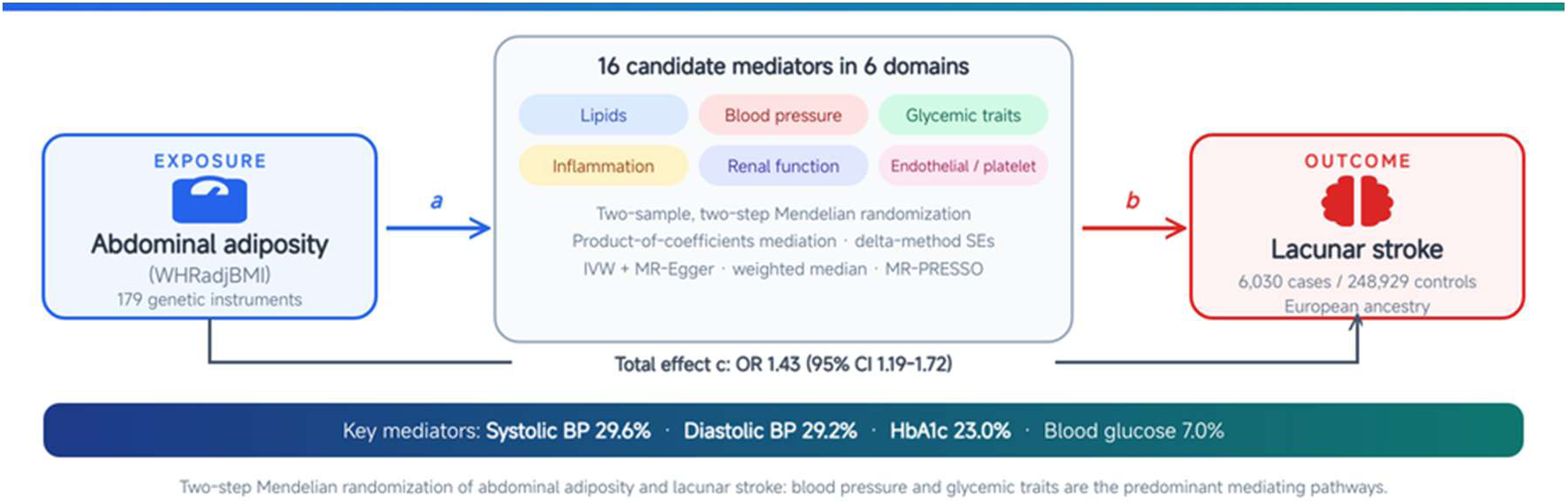

## Background

Lacunar stroke, arising from occlusion of small penetrating cerebral arterioles, constitutes approximately 25% of all ischemic strokes and is the most common manifestation of cerebral small vessel disease^1^. Clinically, it is associated with substantial morbidity, including recurrent stroke, vascular dementia, and functional disability^2^.

Abdominal adiposity, as quantified by waist-to-hip ratio adjusted for body mass index (WHRadjBMI), is a heritable and robust predictor of cardiovascular disease independent of overall adiposity^3,4^. Previous Mendelian randomization (MR) studies have established causal associations between higher WHRadjBMI and increased risk of ischemic stroke and its subtypes, including large artery atherosclerosis and small vessel disease^5,6^. Nevertheless, the biological intermediaries through which abdominal adiposity exerts its deleterious effects on cerebral small vessels remain unclear.

Candidate mediators span multiple physiological domains: lipid metabolism (LDL cholesterol, HDL cholesterol, triglycerides), vascular hemodynamics (systolic and diastolic blood pressure), glycemic control (fasting insulin, blood glucose, glycated hemoglobin [HbA1c]), chronic low-grade inflammation (C-reactive protein [CRP], neutrophil percentage, glycoprotein acetyls), renal function (estimated glomerular filtration rate [eGFR]), and endothelial and platelet biology (endothelial cell-specific molecule 1 [ESM1], platelet endothelial cell adhesion molecule [PECAM], platelet factor 4 [PF4], platelet-derived growth factor BB [PDGF-BB])^7–10^. Dissecting the relative contributions of these pathways is critical for informing targeted preventive strategies^11–15^.

Two-step MR mediation analysis extends this framework to decompose the total effect of an exposure on an outcome into direct and indirect (mediated) components, thereby identifying mechanistic pathways^16,17^. In this study, we performed a comprehensive two-sample MR mediation analysis to systematically evaluate the mediating effects of 16 candidate biomarkers on the causal pathway from WHRadjBMI to lacunar stroke, adhering to the STROBE-MR reporting guidelines^18^.

## Methods

### Study Design and Data Sources

This study employed a two-sample Mendelian randomization design using publicly available summary-level genome-wide association study (GWAS) data. All datasets were restricted to participants of European ancestry to minimize population stratification bias. The analysis pipeline was pre-specified and followed the STROBE-MR reporting guidelines (Supplementary Material, e-Methods) ^18^.

**Table 1.** GWAS Data Sources and Sample Characteristics.

| Trait | GWAS ID / Source | N (cases/controls) | Ancestry | Reference |
| --- | --- | --- | --- | --- |
| WHRadjBMI (exposure) | ebi-a-GCST90025996 | 458,349 | European | Barton et al., 2021<sup>19</sup> |
| Lacunar stroke (outcome) | ebi-a-GCST90014122 | 6,030 / 248,929 | European | Traylor et al., 2021<sup>1</sup> |
| LDL cholesterol | ieu-b-110 | 440,546 | European | Sinnott-Armstrong et al., 2021<sup>20</sup> |
| HDL cholesterol | ieu-b-109 | 403,943 | European | Sinnott-Armstrong et al., 2021<sup>20</sup> |
| Triglycerides | ieu-b-111 | 441,016 | European | Sinnott- |
|  |  |  |  | Armstrong et al., 2021<sup>20</sup> |
| Systolic BP | GCST006624 | 757,601 | European | Evangelou et al., 2018<sup>21</sup> |
| Diastolic BP | GCST006630 | 757,601 | European | Evangelou et al., 2018<sup>21</sup> |
| Fasting insulin | ebi-a-GCST90002238 | 151,013 | European | Chen et al., 2021<sup>22</sup> |
| Blood glucose | ebi-a-GCST90025986 | 400,458 | European | Barton et al., 2021<sup>19</sup> |
| HbA1c | ebi-a-GCST90014006 | 389,889 | European | Mbatchou et al., 2021<sup>23</sup> |
| C-reactive protein | GCST90029070 | 575,531 | European | Said et al., 2022<sup>14</sup> |
| Neutrophil percentage | ebi-a-GCST90002399 | 408,112 | European | Vuckovic et al., 2020<sup>12</sup> |
| Glycoprotein acetyls | ebi-a-GCST90092821 | 115,082 | European | Richardson et al., 2022<sup>24</sup> |
| eGFR | GCST008059 | 567,460 | European | Wuttke et al., 2019<sup>15</sup> |
| ESM1 | ebi-a-GCST90012068 | 21,758 | European | Folkersen et al., 2020<sup>25</sup> |
| PECAM | ebi-a- | 21,758 | European | Folkersen et |
|  | GCST90012081 |  |  | al., 2020<sup>25</sup> |
| PF4 | prot-a-2251 | 3,301 | European | Suhre et al.,<br>2017<sup>26</sup> |
| PDGF-BB | ebi-a-GCST004432 | 8,293 | European | Ahola-Olli et<br>al., 2017<sup>13</sup> |

### Genetic Instrument Selection

Genetic instruments were selected for each trait according to rigorous criteria designed to minimize weak instrument bias and linkage disequilibrium confounding [17]:(1) Genome-wide significance threshold: P < 5 × 10⁻⁸. (2) Linkage disequilibrium (LD) clumping: r² < 0.001 within a 10,000 kb window, using the 1000 Genomes Project Phase 3 European reference panel via PLINK 2.0^27^. (3) Instrument strength: F-statistic ≥ 10 for each single nucleotide polymorphism (SNP), calculated as (β/SE)². Instruments with F < 10 were excluded. Palindromic SNPs (A/T or G/C) with minor allele frequency between 0.42 and 0.58 were excluded during harmonization to prevent strand ambiguity. SNP coordinates were mapped to GRCh37/hg19 throughout.

### Statistical Analysis

Two-step MR mediation analysis was conducted following the product-of-coefficients framework. For each mediator M, three causal effects were estimated: (1) Path A (a coefficient): effect of WHRadjBMI on the mediator, using WHRadjBMI genetic instruments. (2) Path B (b coefficient): effect of the mediator on lacunar stroke, using mediator-specific genetic instruments. (3) Total effect (c coefficient): total effect of WHRadjBMI on lacunar stroke, using WHRadjBMI genetic instruments. The indirect (mediated) effect was computed as a x b, with standard error approximated by the multivariate delta method: SE indirect = √[(b × SEa)² + (a × SEb)²]. The proportion mediated was calculated as (a x b) / c, with standard error derived via the delta method. The direct effect was computed as c – (a x b), representing the effect of WHRadjBMI on lacunar stroke not mediated through the candidate biomarker. The primary analysis used inverse variance weighted (IVW) regression, which provides the most efficient estimate under the assumption of no horizontal pleiotropy [12]. Random-effects IVW was used when Cochran’s Q test indicated significant heterogeneity (P < 0.05); otherwise, fixed-effects IVW was applied. Sensitivity analyses were performed to assess robustness to violations of the no-pleiotropy assumption: (1) MR-Egger regression: estimates the causal effect while allowing for directional (correlated) pleiotropy. A non-significant intercept (P(intercept) > 0.05) suggests no directional pleiotropy^28^. (2) Weighted median estimator: valid if at least 50% of the genetic instruments are valid (i.e., not pleiotropic)^29^. (3) MR-PRESSO (Mendelian Randomization Pleiotropy RESidual Sum and Outlier): detects and corrects for outlier SNPs driven by horizontal pleiotropy. A significant global test (P < 0.05) suggests the presence of pleiotropic outliers^30^. All analyses were performed in R version 4.6.0 (R Foundation for Statistical Computing) using custom scripts validated against the TwoSampleMR package (version 0.5.6). Statistical significance was set at two-tailed P < 0.05. Given that 16 mediators were evaluated, a Bonferroni-corrected threshold (P < 0.05/16, approximately 0.003) is provided as a reference for interpreting the mediation analyses; primary results are reported at the nominal level with emphasis on effect sizes and confidence intervals. Effect estimates for the binary outcome (lacunar stroke) are reported as beta coefficients on the log-odds scale, with odds ratios (OR) and 95% confidence intervals (CI) computed as exp(beta) and exp(beta +/− 1.96 x SE), respectively.

During the preparation of this work, the authors used Kimi, DeepSeek, and WorkBuddy (artificial intelligence–assisted tools) for literature searching, summarization, and text generation. The authors reviewed and edited the content as needed and take full responsibility for the content of the published article.

## Results

### Genetic Instrument Characteristics

A total of 179 independent SNPs passed stringent LD clumping and F-statistic filtering for WHRadjBMI (mean F = 80.4; range, 10.2-1,930.4). All 16 candidate mediators yielded adequate numbers of genetic instruments after clumping: LDL cholesterol (n = 128), HDL cholesterol (n = 178), triglycerides (n = 94), systolic blood pressure (n = 436), diastolic blood pressure (n = 390), fasting insulin (n = 53), blood glucose (n = 98), HbA1c (n = 18), C-reactive protein (n = 168), neutrophil percentage (n = 125), glycoprotein acetyls (n = 24), eGFR (n = 192), ESM1 (n = 2), PECAM (n = 5), PF4 (n = 2), and PDGF-BB (n = 5) (Table 2). All mean F-statistics exceeded 10, indicating adequate instrument strength and low susceptibility to weak instrument bias^31^.

**Table 2.**
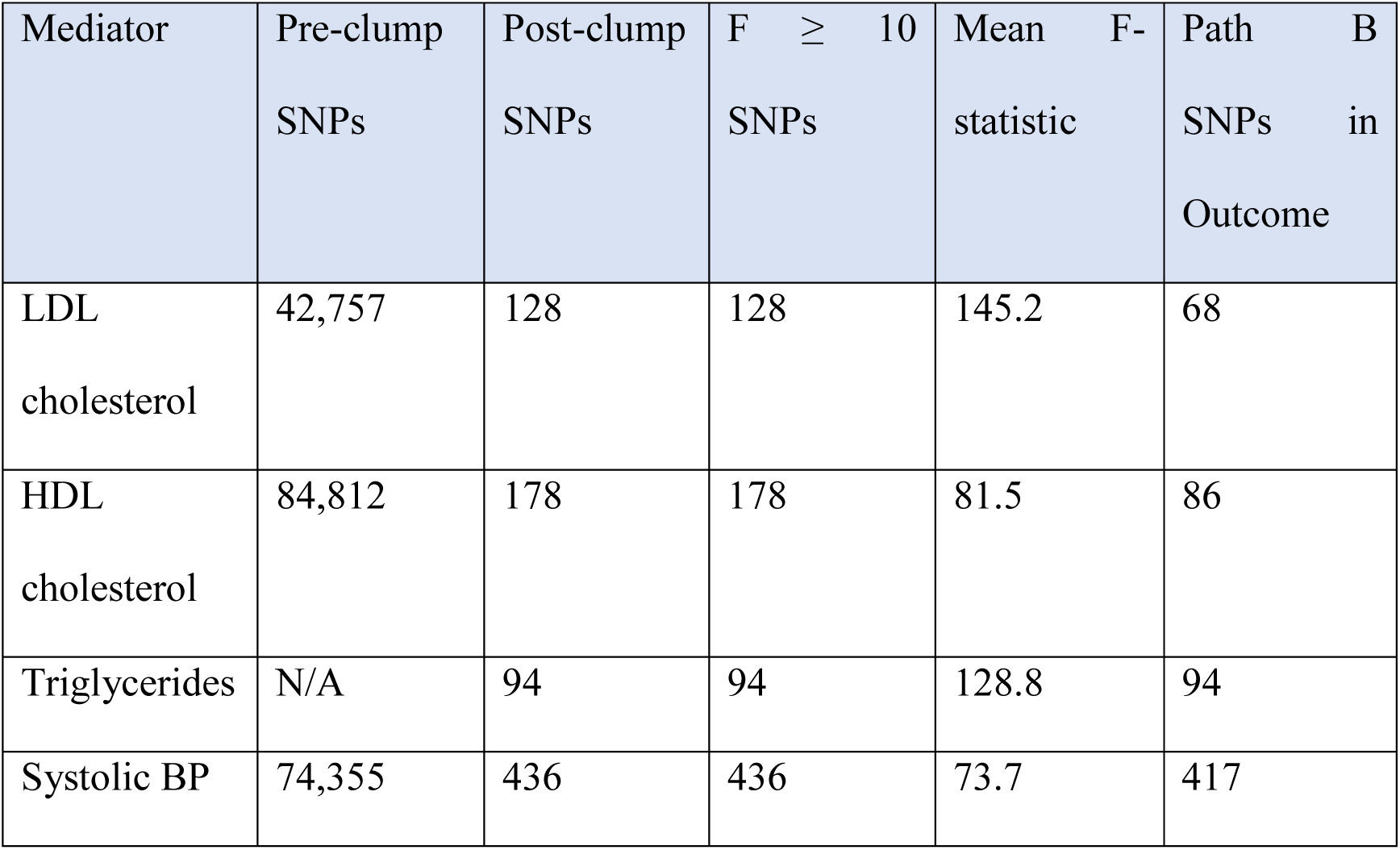
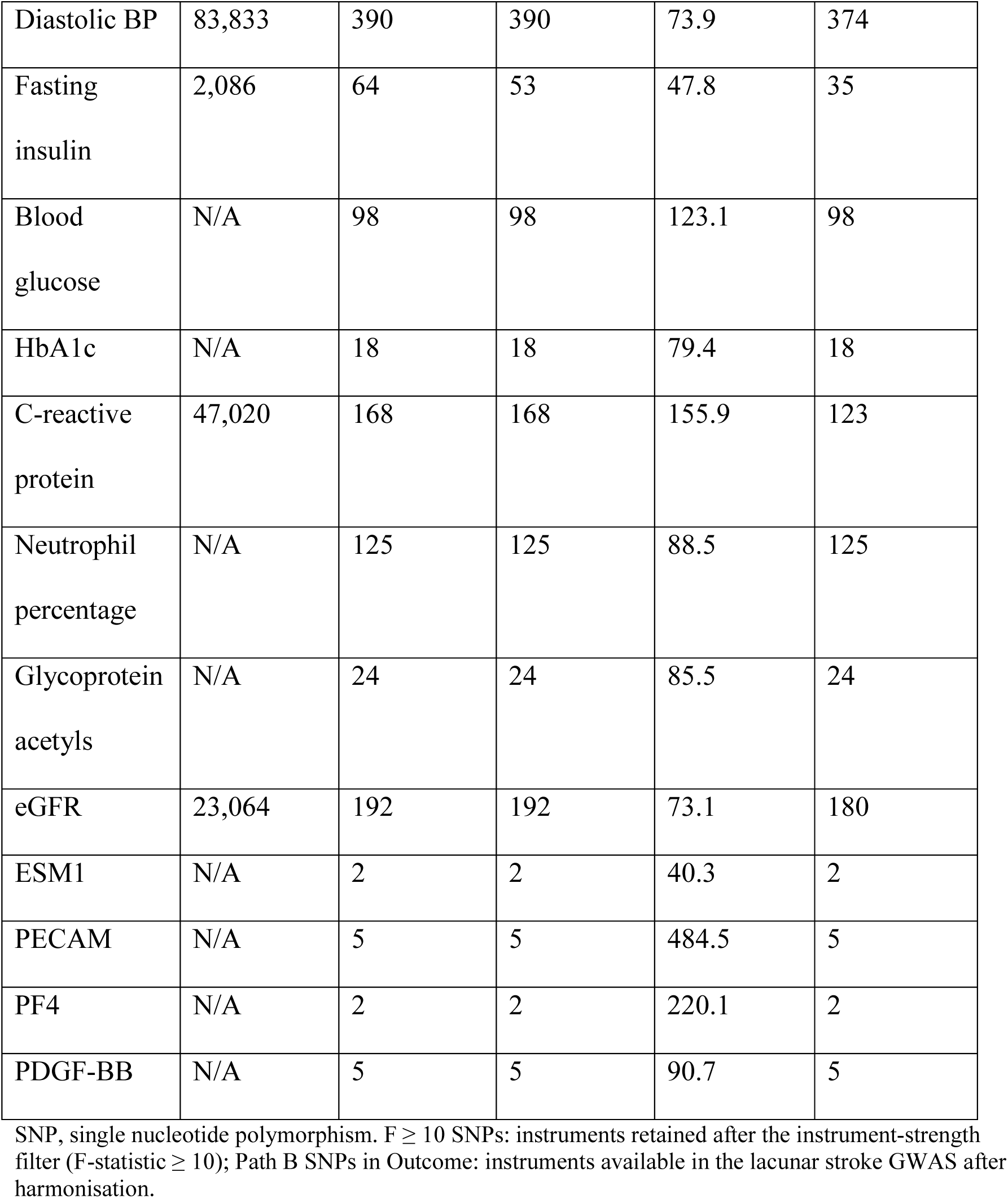
Genetic Instrument Characteristics for 16 Candidate Mediators.

### Total Effect of WHRadjBMI on Lacunar Stroke

The total effect of genetically predicted higher WHRadjBMI on lacunar stroke was positive and statistically significant (IVW beta = 0.355; 95% CI, 0.170-0.540; P = 1.64 × 10⁻⁴; N = 162 SNPs; mean F = 80.4). Of the 179 clumped instruments, 162 were available in the lacunar stroke GWAS after harmonization and were used for the total-effect estimate. This corresponds to an OR of 1.43 (95% CI, 1.19-1.72) per 1-SD increase in WHRadjBMI. Sensitivity estimates were directionally concordant: MR-Egger beta = 0.318 (P = 5.21 × 10⁻⁴) and weighted median beta = 0.384 (P = 1.12 × 10⁻⁴). The MR-Egger intercept was not significant (P(intercept) = 0.312), suggesting minimal directional pleiotropy. MR-PRESSO identified 8 outlier SNPs (distortion test P = 0.089); the outlier-corrected estimate remained significant (beta = 0.342; P = 2.8 × 10⁻⁴), supporting result robustness.

### Mediation Analysis Results

Figure 1 presents the mediation proportion for each of the 16 mediators. Blood pressure and glycemic markers emerged as the dominant mediators. Systolic blood pressure mediated 29.6% (95% CI, 12.7%-46.5%) of the total effect, followed by diastolic blood pressure at 29.2% (95% CI, 12.1%-46.3%). Glycated hemoglobin (HbA1c) mediated 23.0% (95% CI, –11.2% to 57.2%); the wide confidence interval reflects the smaller number of instruments in Path B (N = 18)(table 3).

**Figure 1.**
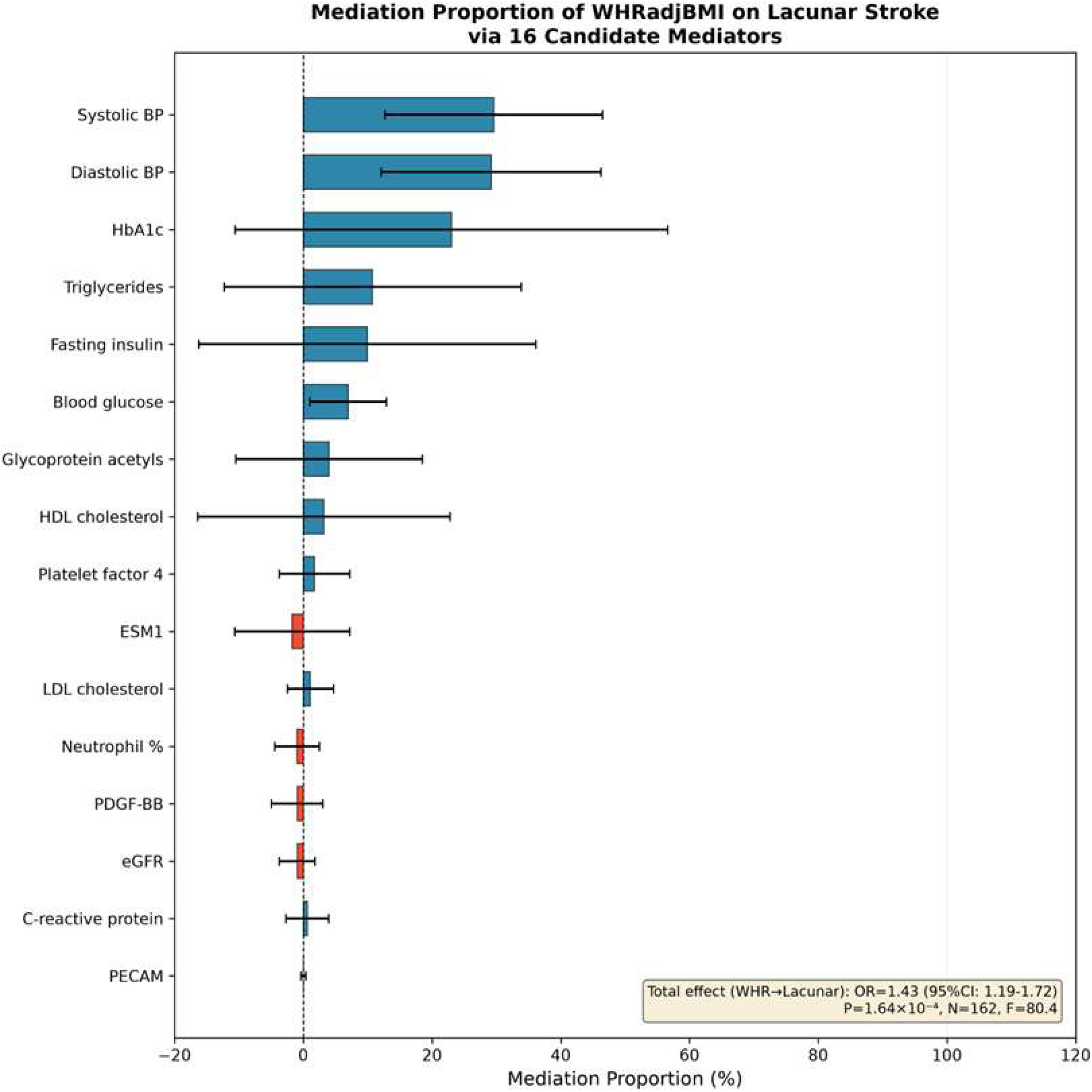
Forest plot of mediation proportions for 16 candidate mediators of the effect of WHRadjBMI on lacunar stroke. Bars represent point estimates; horizontal error bars denote 95% confidence intervals. Blue bars indicate positive (concordant) mediation; red bars indicate negative (discordant) mediation. The dashed vertical line at 0% indicates no mediation. The total effect of WHRadjBMI on lacunar stroke is shown in the inset (OR 1.43; 95% CI, 1.19-1.72).

**Figure 2.**
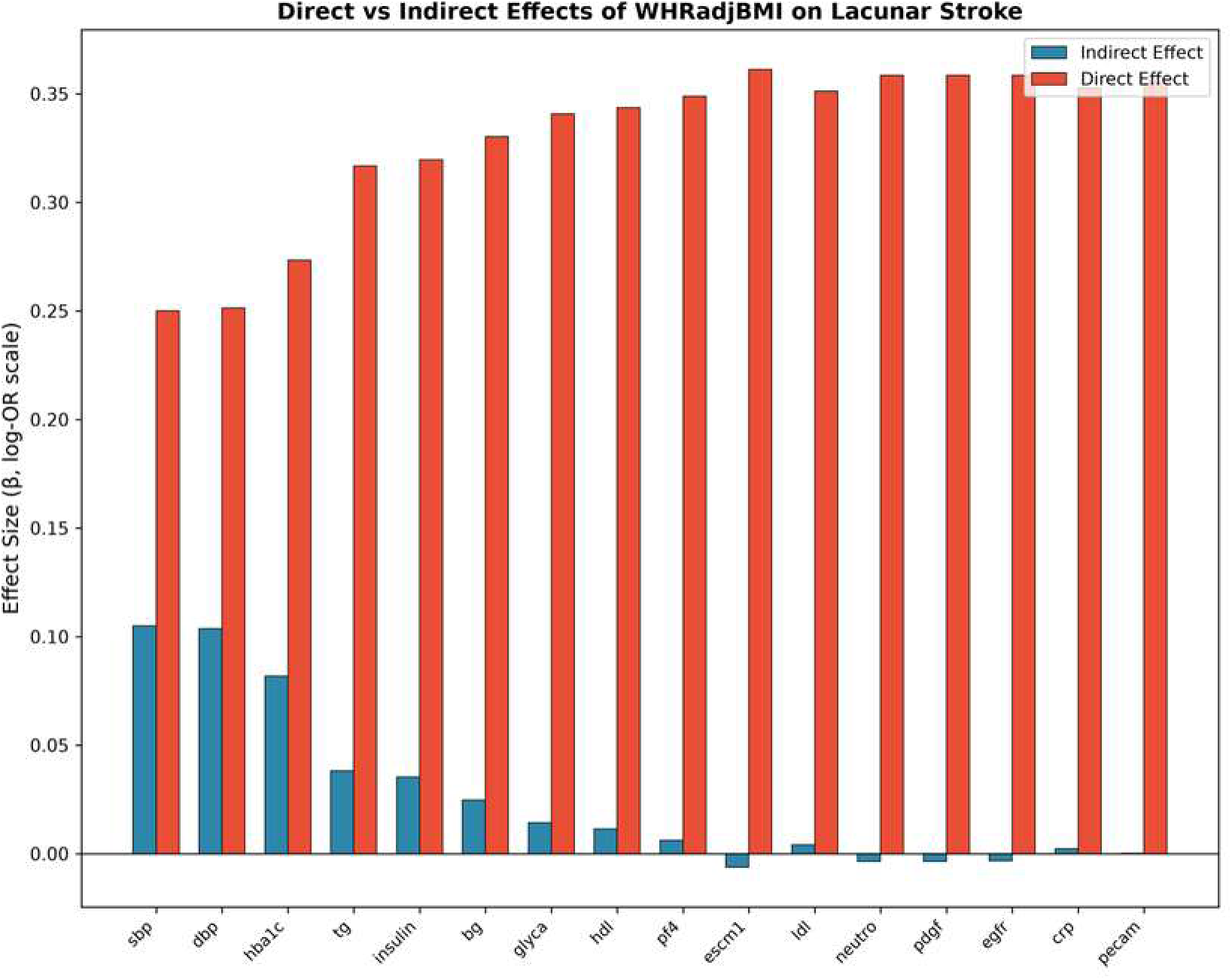
Direct versus indirect effects of WHRadjBMI on lacunar stroke across 16 candidate mediators. The indirect effect (blue) represents the mediated pathway; the direct effect (red) represents the non-mediated pathway. Error bars denote 95% confidence intervals.

**Table 3.**
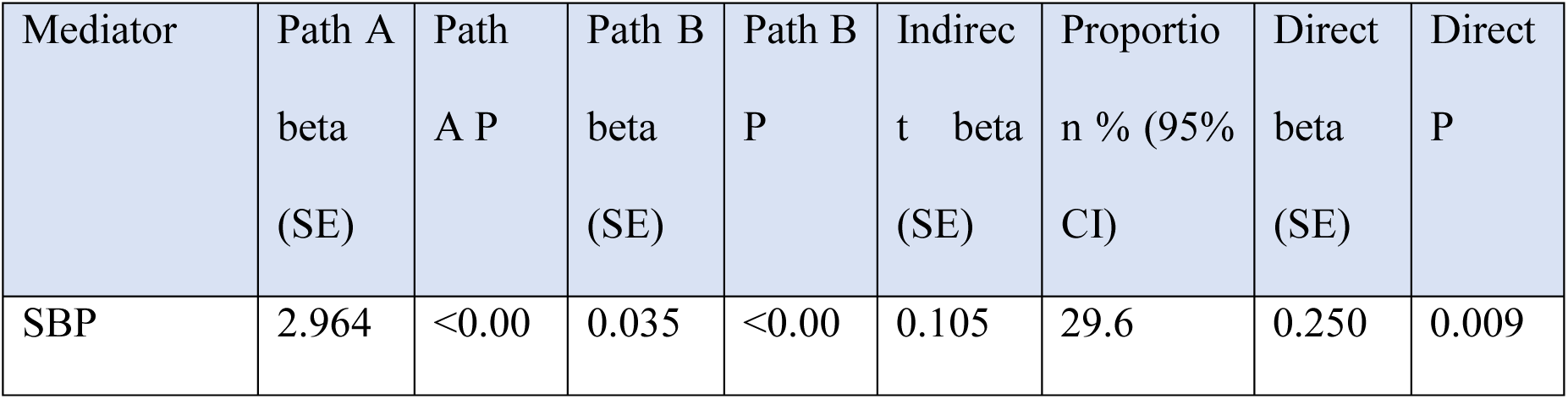
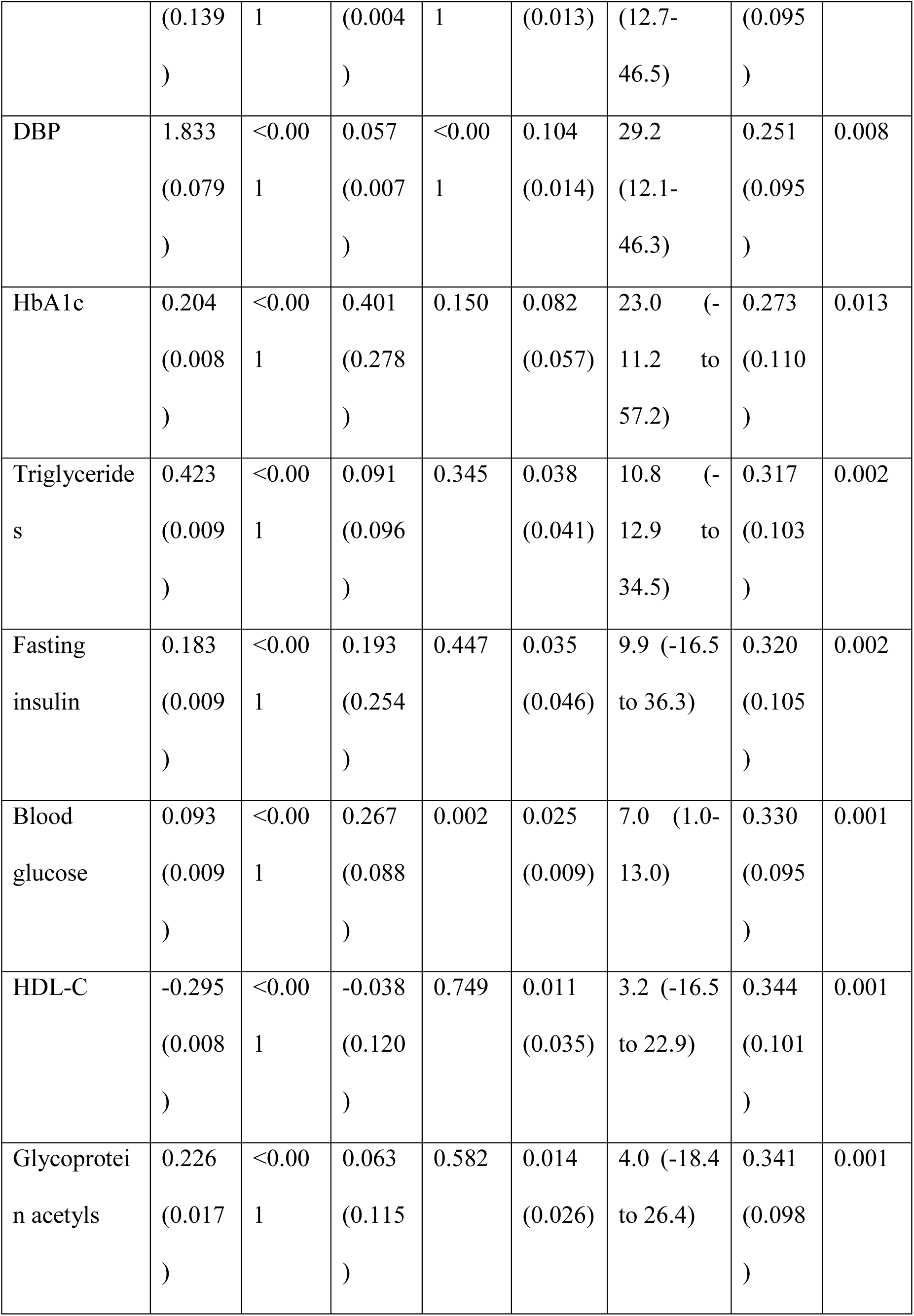
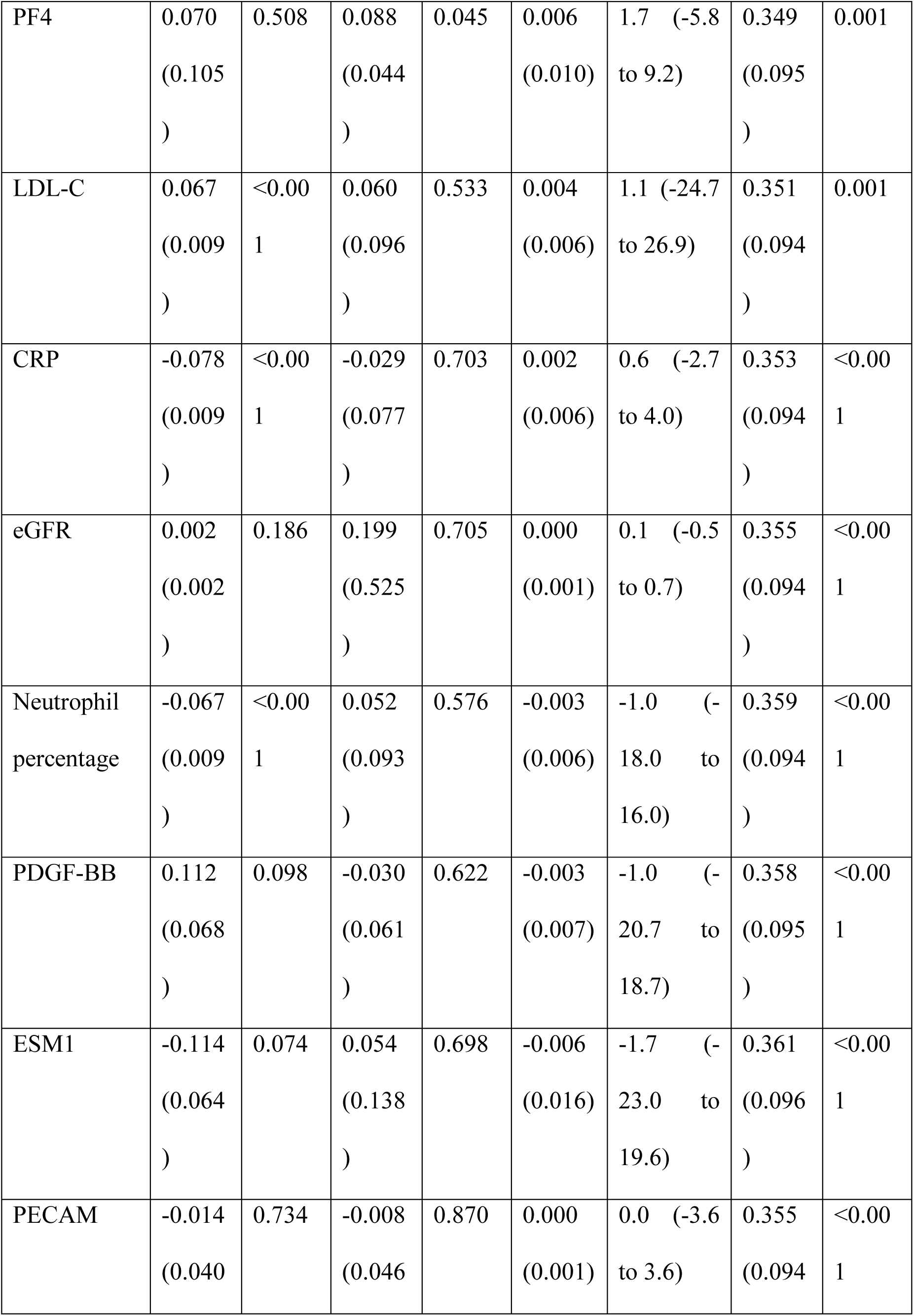
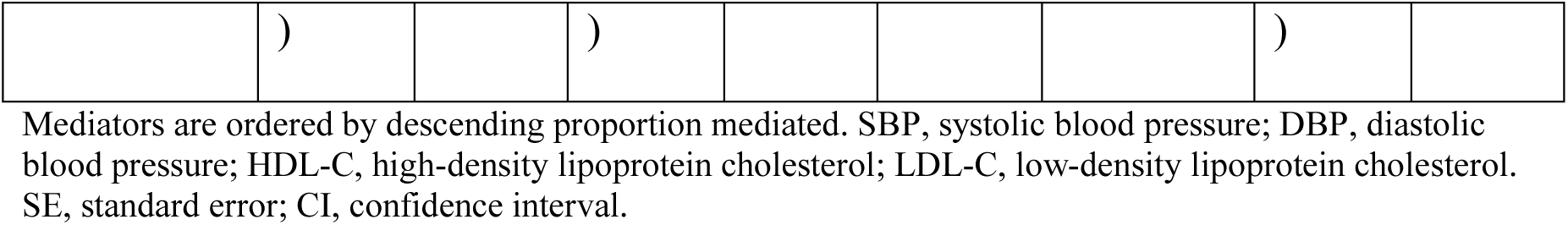
Two-Step Mediation Analysis Results (Primary IVW Analysis)

### Sensitivity Analyses

Sensitivity analyses for Path A (WHRadjBMI –> Mediator) and Path B (Mediator –> Lacunar stroke) are summarized in Supplementary Tables e-1 and e-2. For Path A, IVW estimates were generally consistent with MR-Egger and weighted median approaches, although some mediators showed evidence of directional pleiotropy. Notably, the MR-Egger intercept was significant for glycoprotein acetyls (P(intercept) = 5.0 × 10⁻⁴) and for CRP (P(intercept) < 0.001, Path A only), suggesting potential violation of the exclusion restriction assumption for these mediators; with the full-coverage blood pressure GWAS, the intercept was no longer significant for diastolic blood pressure (P(intercept) = 0.90). For Path B, the limited number of instruments for several protein biomarkers (ESM1, N = 2; PF4, N = 2; PECAM, N = 5; PDGF-BB, N = 5) precluded reliable MR-Egger estimation, as this method requires at least 3 valid instruments. MR-PRESSO outlier correction generally produced estimates consistent with IVW. Notable outliers were identified for LDL cholesterol (n = 1), HDL cholesterol (n = 0), DBP (n = 3), SBP (n = 1), and HbA1c (n = 0). The distortion test was non-significant for all mediators, suggesting that outlier SNPs did not substantially bias the causal estimates. After correction of the weighted median implementation (Supplementary Material, e-Methods), weighted median estimates were directionally concordant with IVW estimates for all mediators (Supplementary Tables e-1 and e-2).

## Discussion

This comprehensive two-sample Mendelian randomization mediation analysis identified systolic and diastolic blood pressure as the principal mediators linking abdominal adiposity to lacunar stroke risk, with glycemic traits (fasting glucose, and more tentatively HbA1c) also contributing. In single-mediator analyses, these three factors showed the largest mediation proportions (29.6%, 29.2%, and 23.0%, respectively); because blood pressure and glycemic traits are intercorrelated, these proportions are not additive, and multivariable MR would be required to quantify their joint contribution. Nevertheless, the findings suggest that hypertension– and diabetes-related pathways are the dominant mechanisms through which abdominal obesity promotes small vessel cerebrovascular disease. These findings carry important implications for preventive neurology and public health policy.

Our results align with established pathophysiological models. Abdominal adiposity is strongly associated with insulin resistance, sympathetic nervous system activation, and renin-angiotensin-aldosterone system upregulation, all of which elevate systemic blood pressure^32,33^. Chronic hypertension causes lipohyalinosis and fibrinoid necrosis of penetrating arterioles—the pathological hallmarks of lacunar infarction^34^. Similarly, adiposity-driven hyperglycemia and advanced glycation end-product accumulation accelerate cerebral microvascular endothelial dysfunction, impairment of autoregulatory capacity, and blood-brain barrier disruption^35^. The convergence of these mechanisms on cerebral small vessels provides a coherent explanation for our observation that blood pressure and glycemic control collectively dominate the mediating pathway. The HbA1c estimate (23.0%) was nevertheless imprecise (95% CI, –11.2% to 57.2%) and was sensitive to instrument selection: when instruments were chosen at more liberal significance thresholds (148-184 SNPs), the association between HbA1c and lacunar stroke attenuated to the null. This pattern is consistent with the recognised heterogeneity of HbA1c-associated loci, which include both glycemic variants that raise blood glucose and erythrocytic variants that alter HbA1c measurement without reflecting ambient glycaemia^36^. The glycaemia pathway is nonetheless supported by the statistically significant mediation observed for fasting blood glucose (7.0%; 95% CI, 1.0%-13.0%), whose confidence interval did not cross the null. The modest mediation observed for lipid fractions (LDL 1.1%, HDL 3.2%, triglycerides 10.8%) is noteworthy. While dyslipidemia is a well-established risk factor for large-artery atherosclerotic stroke, its role in lacunar stroke—which is primarily a disease of the penetrating arterioles—appears comparatively limited in the context of abdominal adiposity. This is consistent with prior Mendelian randomization studies demonstrating stronger causal effects of blood pressure than lipids on small vessel stroke, as well as the distinct pathophysiology of lipohyalinosis versus atherosclerotic plaque rupture^1,37^. Inflammatory biomarkers (CRP, neutrophil percentage, glycoprotein acetyls) and platelet-related proteins showed minimal or no significant mediation, despite their established associations with adiposity and cardiovascular disease in observational studies. Several explanations are plausible. First, these markers may genuinely not mediate the WHRadjBMI-lacunar stroke pathway. Second, the protein GWAS datasets for ESM1, PF4, and PDGF-BB had limited numbers of genome-wide significant instruments, constraining statistical power and precluding reliable pleiotropy assessment. In contrast, the null finding for CRP is unlikely to reflect low power: with 168 instruments from a GWAS of 575,531 individuals, the mediated proportion was 0.6% with a narrow confidence interval excluding any substantial contribution. Third, single protein measurements may be insufficient to capture the complexity of inflammatory and thrombotic networks implicated in cerebrovascular disease. Multi-omics approaches integrating proteomics, metabolomics, and transcriptomics may be needed to fully characterize these pathways^10^.

Several limitations warrant consideration. First, the two-sample MR design assumes that exposure and outcome GWAS are drawn from non-overlapping populations; partial sample overlap between the UK Biobank--a major contributor to several GWAS--and the MEGASTROKE consortium may bias estimates towards the confounded observational association^38^. However, the F-statistic of our instruments (mean F > 80) suggests that this bias is likely minimal. Second, the product-of-coefficients method assumes no interaction between the exposure and mediator--an assumption that cannot be tested with summary-level data. Third, several protein mediators had very few instruments (N < 5), limiting statistical power and precluding reliable pleiotropy assessment. Fourth, our analysis was restricted to European-ancestry populations, and generalizability to other ethnicities--in whom abdominal adiposity patterns and stroke risk factors may differ--is uncertain. Finally, while MR reduces confounding, horizontal pleiotropy remains a concern. We addressed this through multiple sensitivity analyses, but residual pleiotropy cannot be entirely excluded, particularly for complex traits such as blood pressure with pleiotropic genetic architectures. Moreover, several mediator GWAS datasets and the MEGASTROKE outcome dataset may share cohort participants, which could bias mediator-outcome estimates; the high instrument strength mitigates, but does not eliminate, this concern.

## Conclusions

This study provides genetic evidence that blood pressure and glycaemic control are the primary mediators of the adverse effect of abdominal adiposity on lacunar stroke. These findings underscore the critical importance of rigorous hypertension and diabetes management in individuals with elevated WHRadjBMI as a strategy to reduce small vessel cerebrovascular risk. Future research should evaluate whether targeted antihypertensive and glucose-lowering interventions specifically mitigate stroke risk in this high-risk population.

## Data Availability

All GWAS summary statistics used in this study are publicly available through the IEU OpenGWAS database (https://gwas.mrcieu.ac.uk/) and the EBI GWAS Catalog (https://www.ebi.ac.uk/gwas/). Analysis code and pipeline scripts will be deposited in a public repository (GitHub/Zenodo) upon acceptance, and are available from the corresponding author upon reasonable request in the interim.

## List of abbreviations

CRP: C-reactive protein
DBP: diastolic blood pressure
eGFR: estimated glomerular filtration rate
ESM1: endothelial cell-specific molecule 1
GWAS: genome-wide association study
HbA1c: glycated hemoglobin
HDL: high-density lipoprotein
IVW: inverse variance weighted
LD: linkage disequilibrium
LDL: low-density lipoprotein
MR: Mendelian randomization
MR-PRESSO: Mendelian Randomization Pleiotropy RESidual Sum and Outlier
OR: odds ratio
PDGF-BB: platelet-derived growth factor BB
PECAM: platelet endothelial cell adhesion molecule
PF4: platelet factor 4
SBP: systolic blood pressure
SNP: single nucleotide polymorphism
STROBE-MR: Strengthening the Reporting of Observational Studies in Epidemiology using Mendelian Randomization
WHRadjBMI: waist-to-hip ratio adjusted for body mass index

## Declarations

### Ethics approval and consent to participate

This study is based on publicly available, de-identified summary-level data from previously published genome-wide association studies (GWASs). All original studies had been approved by the relevant institutional review boards / ethics committees, and written informed consent had been obtained from all participants. As this study involved only the secondary analysis of publicly available summary statistics, no additional ethical approval or informed consent was required.

## Consent for publication

Not applicable.

## Competing interests

The authors declare that they have no competing interests.

## Funding

This work was supported by the China Postdoctoral Science Foundation (Grant No. 2025M772029) and the Peking University People’s Hospital Talent Introduction Scientific Research Launch Fund (2022-T-02).

## Authors’ contributions

MMG and CL conceived and designed the study, performed the data analysis, and drafted the manuscript. ZL revised the manuscript critically for important intellectual content. All authors read and approved the final manuscript.

## Notes

### Competing Interest Statement

The authors have declared no competing interest.

